# Fibroblast–Neuron Interactions Driving Persistent Pain in Rheumatoid Arthritis (FiND-Pain RA) - a study protocol

**DOI:** 10.1101/2025.01.02.25319882

**Authors:** Mikalena Xenophontos, Friederike C. Baldeweg, Rosie Ross, Zoe Rutter-Locher, Sarah Hill, Sarah Ryan, Mosab Ali Awadelkareem, Shing T. Law, David L. Bennett, Christopher D. Buckley, Frances Humby, Bruce W. Kirkham, Franziska Denk, Leonie S. Taams

**Affiliations:** Kennedy Institute of Rheumatology, University of Oxford, UK; Nuffield Department of Orthopaedics, Rheumatology and Musculoskeletal Sciences, University of Oxford; Centre for Inflammation Biology and Cancer Immunology (CIBCI), Department of Inflammation Biology, School of Immunology & Microbial Sciences, King’s College London, London, UK; Department of Rheumatology, Guy’s and St Thomas’ NHS Foundation Trust, London, UK; Nuffield Department of Clinical Neurosciences, University of Oxford, UK; Wolfson Sensory, Pain and Regeneration Centre (SPaRC), School of Neuroscience, King’s College London, London, UK

**Author notes:** Mikalena Xenophontos and Friederike Baldeweg are co-first authors and contributed equally to the manuscript. Franziska Denk and Leonie Taams are co-senior authors and contributed equally to the manuscript. A PUMIA *(Pain Phenotypes and their Underlying Mechanisms in Inflammatory Arthritis)* sub-study. **Corresponding author** - Friederike C. Baldeweg –, Centre for Inflammation Biology and Cancer Immunology (CIBCI), Kings College London, SE1 9NH.

## Abstract

**Introduction:** Pain in patients with rheumatoid arthritis (RA) is an unmet clinical need. Targeting joint inflammation with disease modifying antirheumatic drugs (DMARDs) has not resulted in the anticipated reduction in pain for many patients. This can partly be explained by the concept of central sensitisation whereby spinal and supraspinal pathways have a lower threshold of activation leading to increased perception of pain. Synovial stromal cells such as fibroblasts are also thought to play a role through peripheral sensitisation of nerves in the joint. Synovial fibroblasts are known to produce pro-algesic mediators such as interleukin 6 (IL-6) and nerve growth factor (NGF) at the mRNA level. These pro-algesic mediators could activate sensory nerve fibres that send signals from the joint to the spinal cord, thereby driving persistent pain in RA. The purpose of this study is to evaluate which pro-algesic mediators are produced by lining versus sublining fibroblasts and whether the level of these mediators correlates with clinical measures of pain in patients with RA.

**Methods and analysis:** FiND-Pain RA is a multi-centre observational study which will recruit 50 patients with seropositive RA who attend the rheumatology department of Guy’s and St Thomas’ Hospital, London and the Nuffield Orthopaedic Centre, Oxford. Clinical examination, pain-focused patient reported outcome measures (PROMs), ultrasound examination and ultrasound guided synovial biopsy of the knee will be performed. The levels of known and putative pro-algesic mediators will be measured in fibroblasts from the lining and sublining layer of the synovium. The location and spatial morphology of sensory nerve fibres and their proximity to lining and sub-lining fibroblasts will be characterized. The primary outcome will be to determine whether the knee pain scores of participants correlate with the level of LIF, a novel putative pain-mediator expressed in sub-lining fibroblasts. The secondary outcomes will be to determine whether other pro-algesic mediators produced by lining or sub-lining fibroblasts correlate with clinical measures of pain and to assess the location and proximity of sensory nerve fibres to lining versus sub-lining fibroblasts.

**Ethics and dissemination:** The study has been approved by the Bromley Research Ethics Committee (REC: 21/LO/0712). The findings of this study will be disseminated through open-access publications, as well as scientific and clinical conferences.

## Introduction

### Background

Rheumatoid arthritis (RA) is an immune mediated inflammatory disease which causes pain, joint damage, and subsequent disability ^1 2^. Pain has been ranked as the highest priority for improvement by patients with RA, yet remains an unmet clinical need ^3^. The use of biological and targeted therapies has reduced joint inflammation and damage, however, has not improved pain to the same extent ^4 5^. 75% of European and 82% of US patients reported moderate to severe persistent pain even though they felt that their RA was ‘somewhat to completely’ controlled ^6^.

Traditionally, pain has been attributed to joint inflammation in patients with RA. Sensory nerve fibres are known to be present in the deeper layers of the synovium in RA, now commonly referred to as sub-lining ^7 8^. Pro-inflammatory cytokines in the joint, such as IL-6 and TNF-alpha, have their corresponding receptors on these sensory nerve fibres and are thought to initiate noxious signalling which is relayed via nociceptive pathways in the spinal cord to the brain where they can be perceived as pain ^9^. However, given that pain frequently does not resolve fully with biological therapies targeting such cytokines in RA, there must be alternative drivers of pain which are uncoupled from ‘classical’ synovial tissue inflammation ^10 11^.

Central sensitisation is widely accepted by rheumatologists as a mechanism whereby pain is potentiated and amplified through the phenomenon of neurons within central nervous system nociceptive pathways responding more readily to ordinary levels of sensory neuron input ^12^. Fibromyalgia is a condition thought to be caused by central sensitisation and is common amongst patients with moderate to severe RA, both in the presence and absence of inflammation ^13^. However, fibromyalgia is not the sole explanation for persistent pain in the absence of traditional measures of joint inflammation. The TITRATE-US study recently described that 27% of patients with moderate to severe RA did not meet the American College of Rheumatology (ACR) 2010 criteria for fibromyalgia and displayed no evidence of joint inflammation, as measured by ultrasound examination of the joints ^13^. This points to alternate mechanisms driving persistent pain in RA.

One potential option is the presence of inflammation caused by dysregulation of tissue resident cells, such as fibroblasts, which are well-known for driving and maintaining joint pathology in RA ^14-16^. Recent work suggests that fibroblasts may also be involved in the generation of pain. For example, synovial fibroblasts activated by TNF-alpha can promote neuronal sensitisation in an in vitro model ^17^. Furthermore, in a clinical setting, it has been reported that a gene expression module, consisting of 815 genes over-represented in lining layer fibroblasts, correlates with pain in the presence of low inflammation in RA synovial biopsy samples ^18^. Finally, our group has re-examined publicly available single cell transcriptomic data from synovial biopsies from individuals with RA ^19-21^. This analysis demonstrated that fibroblasts in the sub-lining layer of the synovium are particularly enriched for both known and putative pro-algesic mediators such as nerve growth factor (NGF) ^22^, interleukin 6 (IL-6) ^23^, complement C3 ^24^, leukaemia inhibitory factor (LIF) ^25^ and IL-11 ^26^. C3, LIF and IL-11 are considered putative pro-algesic mediators based on mRNA expression of their receptors (C3AR1, CR2, LIFR and IL11RA) on sensory neurons and prior literature ^24 25^. Moreover, we have recently shown that both LIF and IL-11 can induce intracellular activation of human stem-cell derived sensory neurons ^26^.

Intriguingly, in RA, there appears to be an expansion of perivascular sub-lining THY1+ fibroblasts and a concomitant retraction of sensory nerve fibres from the lining into the sub-lining layer ^7 27^. It is thus feasible that nerve fibresare attracted to move back into the sub-lining by perivascular and fibroblast-derived neurotrophic mediators such as NGF (Figure 1). Altogether, the currently available evidence leads us to hypothesize that fibroblasts, and possibly other stromal cells, in the synovium of patients with RA are key drivers of persistent pain.

**Figure 1.**
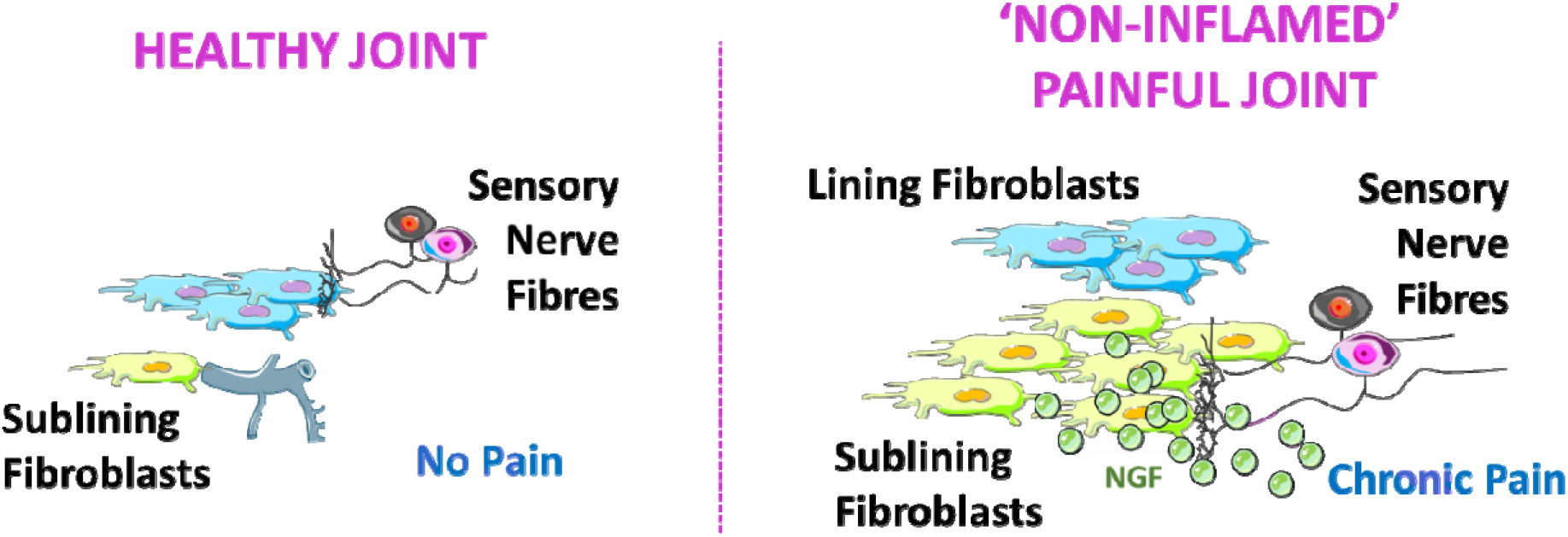
Schematic representation of how perivascular sub-lining fibroblasts may be driving pain in rheumatoid arthritis. Pro**-**algesic mediators such as nerve growth factor produced by sub-lining fibroblasts attract sensory nerve fibres to the sub-lining layer. Sensory nerve fibres become activated and signal via the nociceptive pathways to the spinal cord and brain where pain is perceived. This figure was in part compiled using images from Servier Medical Art under a CC-BY-4.0 licence.

### Objectives

FiND-Pain RA aims to understand whether synovial fibroblasts produce pro-algesic mediator which could be driving pain in RA.

### Methods and Study Design

This is a multicentre observational study which will recruit 50 participants with established seropositive RA who meet the ACR/EULAR 2010 criteria; we will assess their disease activity and phenotype their pain. Clinical examination, pain-focused patient reported outcome measures (PROMs), ultrasound examination and ultrasound guided synovial biopsy of the knee will be carried out. In the laboratory, synovial tissue biopsies will be digested, and sublining and lining fibroblasts will be sorted using fluorescence activated cell sorting (FACS). RNA and protein analyses will be used to measure the levels of pro-algesic mediators within these two cell types and correlate them with clinical measures of pain. Sensory neurons within the synovial tissue will be imaged to examine their location and proximity to sub-lining fibroblasts. *In vitro* assays such as neurite outgrowth assays and micro-electrode arrays (MEA) will be used to assess the ability of synovial fibroblasts to sensitise human induced pluripotent stem cell (hiPSC)-derived sensory neurons.

#### a) Setting

The study will recruit at two medical centres: Guy’s and St Thomas’ Hospital in London and Nuffield Orthopaedic Centre in Oxford. Patient recruitment will commence in September 2024 and end in September 2026. Patients will be identified by their direct care team through Rheumatology outpatient clinics or through review of electronic medical data.

#### b) Participants

Clinicians or research nurses who are trained in Good Clinical Practice (GCP) will discuss the study with potential participants at their Rheumatology clinic appointment and provide them with a Participant Information Sheet (PIS). Alternatively, if potential participants do not have a clinic appointment in the near future, they can be called by the chief investigator or study delegates, and a PIS can be sent by email or posted to the participants if they show an interest in the study. Participants will be given at least 24 hours to review the PIS before they are contacted for consent. Willing study participants will attend an appointment at a convenient date and time, where written consent will be obtained. Alternatively, consent can be gained virtually, and participants can email the signed consent form to the researcher. Written consent will be obtained after the nature of the study, including potential risks and benefits, are explained to the participants, and the participants have had the opportunity to ask any questions. All participants will retain a copy of their signed consent form, while the original will be kept in the site file.

#### c) Sample size and rationale

For this study, 50 patients with seropositive rheumatoid arthritis will be recruited (approximately 25 patients at each site). With n = 50, we will have an 80% chance to detect a Spearman’s correlation (LIF vs. average knee pain) of 0.36 or larger with alpha = 0.05. In case of under-recruitment, n = 40 still has an 80% chance to detect two correlations of rho = 0.41 or larger at alpha = 0.05. In both instances, we will be conducting a one-sided analysis assuming that pain will increase as mediator levels increase.

#### d) Eligibility criteria (inclusion and exclusion)

Subject inclusion criteria:

1. Diagnosis of seropositive rheumatoid arthritis (rheumatoid factor and/or cyclic citrullinated peptide (CCP) positive) meeting 2010 ACR/EULAR classification criteria
2. Reporting a knee pain numerical rating scale (NRS) ≥3 on a 0-10 scale; we will aim to recruit study participants that span a range of knee pain scores
3. Symptom duration ≥24 months
4. Stable dose of conventional synthetic DMARD therapy for ≥4 weeks or, if taking steroids, on prednisolone dose ≤10 mg/day or equivalent

<u>Subject exclusion criteria:</u>

1. Under 18 years of age
2. Unable or unwilling to provide informed written consent
3. Unable to comply with study protocols
4. Pregnant or breastfeeding
5. Current or recent (last 90 days) treatment with investigational agents
6. Knee osteoarthritis ≥grade 2 on the Kellgren and Lawrence classification system on a previous knee X-ray held on the Trust’s medical imaging platform
7. Diagnosis of fibromyalgia based on ACR 2016 criteria
8. Treatment with biologic therapy for rheumatoid arthritis or immunosuppressant therapy other than for inflammatory arthritis
9. Use of intra-articular steroids in the affected knee or intramuscular steroids within the last 4 weeks
10. Regular use of direct oral anticoagulant therapy (DOAC), warfarin, high dose Aspirin/Clopidogrel ≥150 mg/day or patients with bleeding diathesis
11. Diagnosis of other painful condition that could act as a confounding variable e.g. peripheral neuropathy
12. Use of non-steroidal anti-inflammatories, opioids, gabapentin, or pregabalin within 24 hours before assessment

### Data collection procedures

#### a) Clinical data collection and pain phenotyping

Demographic and clinical data will be obtained using history and review of electronic records including age, gender, ethnicity, rheumatoid factor, cyclic citrullinated peptide antibody status, comorbidities, medication, inflammatory markers (within the last 28 days), extra-articular features of rheumatoid arthritis, smoking status, body mass index and employment status.

A knee pain score using a visual analogue scale will be obtained from both knees on three occasions prior to synovial tissue sampling; an average score per knee will be taken forward for analysis. Study participants will be asked the following question for each knee: *How would you rate your pain on a 0-10 scale in your (left/right) knee, over the last 24 hours, where 0 is “no pain” and 10 is “pain as bad as could be”?* Participants with a knee pain score between 3 (mild pain) and 10 (severe pain) will be recruited to the study.

As part of their clinical pain phenotyping, participants will complete the short form McGill questionnaire, the PainDETECT questionnaire, and the Intermittent and Constant Osteoarthritis Pain (ICOAP) questionnaire. The short form McGill questionnaire is a validated tool used to assess both the quality and intensity of pain^28^. The PainDETECT questionnaire is a validated questionnaire in rheumatoid arthritis which will be used to assess for the presence of neuropathic-like pain^29^. The ICOAP questionnaire will be included to allow comparison to pain data collected from the Restoring Joint Health and Function to Reduce Pain (RE-JOIN) consortium, part of a National Institute of Health effort to end the opioid public health crisis in the United States^30^. Additional questionnaires will be used to assess PROMs of anxiety, depression and quality of life including the Patient Health Questionnaire-Somatic, Anxiety and Depressive Symptoms (PHQ-SADS), the Musculoskeletal Health Questionnaire (MSK-HQ) and the EuroQol EQ-5D-5L, respectively.

As part of our assessment of pain perception, we will use pressure algometry as a measure of central and peripheral sensitisation in our participants. Pain pressure thresholds (i.e. the pressure at which the stimulus first becomes painful) will be measured over the medial aspect of the knee (peripheral sensitisation) and at the trapezius (additional central sensitisation) bilaterally. A commercial automated pressure algometer will be applied at a steady rate (approximately 0.5kPa/second) and the participant will be asked to report ‘as soon as the usual sensation of pressure changes towards an additional sensation of burning, stinging, drilling or aching’ at which point the test will stop and the reading will be recorded.

#### b) Disease activity scoring

Disease Activity Score of 28 joints (DAS28CRP) will be used to assess disease activity.

#### c) Peripheral blood sampling

Blood tests for C-reactive protein (CRP) will be taken unless CRP data from the last 28 days are already available. In addition, up to 50 ml of blood will be taken for research, provided the participant has given consent. Blood will be transferred to the laboratory where peripheral blood mononuclear cells will be isolated (see protocol below).

#### d) Ultrasound examination of the knee

Musculoskeletal ultrasound will be performed using a LOGIQ E10 ultrasound machine with a 10-14MHz linear transducer at GSTT and a LOGIQ E9 ultrasound machine with a GE ML6-15 transducer at Oxford. Ultrasound images will be taken of the knee from the supra-patella pouch midline, and medial and lateral joint lines. Grey scale (GS) images for synovial thickness and power doppler (PD) images for synovial vascularity will be taken. The PD settings will be adjusted to the lowest permissible pulse repetition frequency to maximise sensitivity and to the maximum colour gain required not to create artefactual noise. The grade of synovial hypertrophy on grey scale and power doppler will be recorded, and the knee with the highest grade of inflammation as defined by the standardised OMERACT scoring system will be biopsied ^31^.

#### e) X-ray

Knee X-rays taken as part of standard clinical care will be scored using the Kellgren and Lawrence classification system, and patients with a grade ≥2 will be excluded from the study (refer to exclusion criteria). No additional X-ray will be performed as part of this research study.

#### f) Synovial biopsy sampling

The ultrasound guided biopsies will be performed in a clean procedure room using sterile aseptic technique by suitably trained operators.

Under ultrasound guidance, local anaesthetic will be injected using a 23G needle into the subcutaneous tissue. Using a 19G/21G needle, any fluid will be aspirated and stored in Na-Hep tubes for research purposes with patient consent. Subsequently 1% lidocaine will be injected inside the joint. A Quick-Core Biopsy Needle (16/14G) will be placed within the joint capsule under ultrasound guidance. The maximum tolerated number of biopsies will be taken, with up to 20 biopsies per knee. A sterile dressing will be applied over the biopsy site.

12 of the biopsies will be placed into a cryo-vial containing 2 ml of CryoStor® CS10 and transferred to the Centre for Inflammation Biology and Cancer Immunology for cryopreservation in liquid nitrogen as described in ^32^. 8 biopsies will be placed into periodate-lysine-paraformaldehyde (PLP) buffer (see laboratory techniques) and transferred to the Kennedy Institute of Rheumatology, Oxford. In the case that less than 20 biopsies are collected, biopsies will be prioritised for storage in CryoStor® CS10.

Participants will be contacted by telephone within 72 hours of the procedure to monitor for adverse events. If an adverse event is suspected, e.g. septic arthritis or haemoarthrosis, the patient will be reviewed in person in the rheumatology clinic in a timely manner and appropriate management procedures followed.

### Synovial tissue digestion and cell sorting

The cryopreserved synovial tissue biopsies will be thawed by warming the tube quickly at 37°C in a water bath and transferring the contents immediately into a fresh tube containing digestion buffer (5 ml serum free RPMI, 300 µg/ml Liberase TL, 100 µg/ml DNase I). The tube will be placed horizontally into a shaking incubator set at 37°C and 200 rpm for 1 hour and 30 minutes. After the digestion, the contents of the tube will be passed through a 70 µM strainer and washed with 10 ml RPMI containing 10% fetal bovine serum (FBS), after rinsing the digestion tube first to collect any remaining cells. The cell solution will then be centrifuged at 220 x g for 10 minutes.

After centrifugation, the supernatant will be discarded, and the cell pellet resuspended in 1 ml PBS and transferred into a 1.5 ml tube. The tube will be centrifuged at 5900 x g, 4°C for 3.5 minutes and subsequently blocked using human serum albumin for 10 minutes at room temperature. After blocking, the cells will be washed via centrifugation with PBS and resuspended in the staining mix and incubated at 4°C for 30 minutes. The staining mix will include antibodies for the following markers: CD90 (marker for sub-lining fibroblasts), PDPN (pan-fibroblast marker), CD45 (immune cell marker), CD146 (mural cell marker), CD31 (endothelial cell marker), CD235a (red blood cell marker), CD3 (T cell marker) and CD14 (myeloid cell marker), in addition to a Live/Dead dye.

After staining, the cells will be washed via centrifugation with cell sorting buffer (PBS with 0.5% BSA, 2 mM EDTA). Finally, the cells will be resuspended in cell sorting buffer and put on ice ready for FACS.

To perform FACS, a CytoFlex SRT cell sorter will be used following the gating strategy depicted in **Figure 2**. PDPN+CD90+ sub-lining and PDPN+CD90-lining fibroblasts will be sorted for both RNA and protein analysis. Additionally, T cells (CD45+CD3+) and myeloid cells (CD45+CD14+) will be sorted for RNA analysis only.

**Figure 2.**
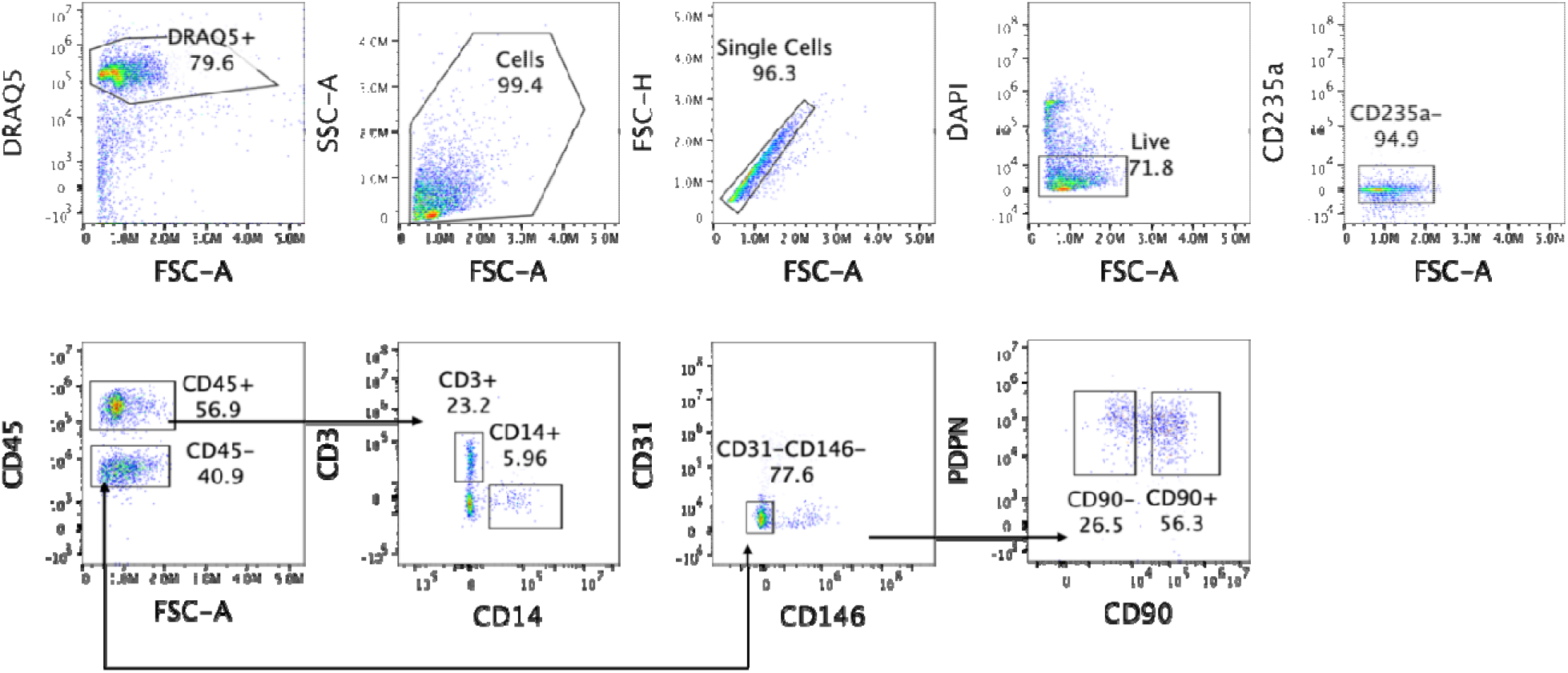
Gating strategy for sorting of PDPN+CD90+ sublining and PDPN+CD90-lining fibroblasts. DRAQ5 is used to distinguish between cells and debris (1^st^ plot), after which cells are further selected based on their forward and side scatter (2^nd^ plot). Single cells are gated (3^rd^ plot) followed by gating for DAPI negative (= live) and CD235a-cells (4^th^ plot). Immune cells will then be identified using CD45 (5^th^ plot), followed by gating on CD3+ T cells and CD14+ myeloid cells (6^th^ and 7^th^ plots). From the CD45-cells, endothelial and mural cells will be excluded using CD31 and CD146 (8^th^ plot). The remaining cells are then gated for the two fibroblast (PDPN+) subsets based on their CD90 expression.

### Synovial tissue confocal imaging

Imaging of RA biopsies will be carried out at the Kennedy Institute of Rheumatology, University of Oxford. Eight RA synovial biopsies will be fixed in Periodate Lysine Paraformaldehyde (PLP) buffer for 24 hours at room temperature, washed 3 times in sterile PBS (Gibco) before being cryoprotected in 15% sucrose (Scientific Laboratory Supplies) for 24 hours and 30% sucrose for up to 5 days. Following cryoprotection, the slides will be washed 3 times in PBS before being embedded and frozen in Optimal Cutting Temperature compound (OCT Tissue-Tek). 50 µm and 10 µm sections will be cut onto slides coated in chromium potassium sulphate (Sigma-Aldrich) and Gelatin (type A, porcine skin, Sigma-Aldrich) using a Leica CM3050 S Cryostat (Leica Biosystems) and left to dry for 24 hours at room temperature. Slides will be rehydrated in PBS 3 times for 5 minutes. Hematoxilin and eosin staining will be completed on the 10 µm sections using the Histology Facilities at the Kennedy Institute for Rheumatology. The 50 µm sections will be used for immunofluorescence staining; first, they will be incubated in 1% SDS solution for 5 minutes as a brief antigen retrieval step, before being washed 3 times in PBS. The slides will then be blocked in a buffer consisting of 5% donkey serum (Bio-Rad), human Fc block (Miltenyi) and 1X Perm/Wash Buffer (BD Biosciences) for 1 hour at room temperature. Slides will then be washed for 5 minutes in PBS before being stained with DAPI for 15 minutes. The slides will then be washed once again in PBS and incubated with primary antibodies (diluted in 3% BSA in PBS) overnight at 4°C. Primary antibodies include and are not limited to the following: CD31 (mouse, clone WM59, Biolegend), PGP9.5,rabbit polyclonal, Zytomed), Calcitonin Gene Related Peptide (CGRP, sheep, Enzo). They will be applied onto the slides in antibody diluent (3% BSA in PBS). The slides with then be washed in PBS and incubated with secondary antibodies at room temperature for 1 hour. Secondary antibodies include donkey anti mouse Alexa Fluor 488 (Abcam, ab150153), donkey anti rabbit 555 (Abcam, ab150074) and donkey anti sheep 647 (Abcam, ab150179). Slides will then be mounted using ProLong Gold Antifade Mountant (Thermo Fisher). Images will be obtained using a Zeiss LSM 980 confocal microscope.

### Peripheral blood or synovial fluid mononuclear cell (PBMC or SFMC) isolation and cryopreservation

Patient-matched blood and synovial fluid (if available) will be taken for isolation and cryopreservation of immune cells, as well as for collection of cell-free serum and synovial fluid. PBMCs will be isolated from whole blood using the density gradient centrifugation technique. Initially, blood will be diluted 1:1 with sterile PBS and then carefully layered on top of Lymphoprep™ using SepMate tubes (25 ml per tube). The tubes will be centrifuged at 1200 x g for 10 minutes, with the brakes off. The PBMCs are then collected from the interphase and washed via centrifugation with PBS. After washing, the cells are counted and cryopreserved at a density of between 10 million and 20 million cells in 1 ml freezing media (RPMI, FBS and DMSO) in a liquid nitrogen tank.

The serum tube will be left for at least 30 minutes to ensure the blood has sufficiently clotted. It is then spun at 2000 x g for 10 minutes, after which five aliquots are taken, between 0.2 ml and 1 ml depending on the volume of serum available. The serum aliquots will be stored at −80°C.

Before isolating SFMCs, two 1 ml aliquots of the synovial fluid will be taken and spun at 6000 x g for 3.5 minutes. The cell free fluid above the pellet will be aliquotted and stored at −80°C. The remaining synovial fluid will be transferred into a 180 mL sample pot and diluted as required with PBS. The isolation and cryopreservation of cells will then be the same as for PBMC isolation.

## Data Analysis

### Primary and secondary outcome measures

The primary outcome of FiND-Pain RA is to determine whether the average pain score of the biopsied knee correlates with the level of LIF, a putative pain-mediator in sub-lining fibroblasts.

The secondary outcomes of FiND-Pain RA are to determine whether 1) sub-lining fibroblasts generally produce more pro-algesic mediators compared to lining fibroblasts and whether this is correlated with the average knee pain score of participants; 2) the ratio of sub-lining to lining fibroblasts is higher in individuals who have higher average knee pain scores; 3) there are more sensory nerve fibres in close proximity to sub-lining versus lining fibroblasts; 4) sub-lining fibroblasts (or their mediators) obtained from participants with higher knee pain scores have greater effect on the outgrowth and excitability of hiPSC sensory neurones using neurite outgrowth assays and MEAs respectively; 5) the levels of pro-algesic mediators correlate with other clinical pain data such as painDETECT scores and pressure algometry.

### Ethics and dissemination

The study has been approved by the Bromley Research Ethics Committee (REC: 21/LO/0712). Results will be disseminated through open-access publications, scientific and clinical conferences, and in partnership with patient involvement groups.

### Trial status

The current protocol version is 4.0 (09/02/2023). Recruitment is expected to begin September 2024 and end approximately September 2026.

## Data Availability

All data produced in the present study are available upon reasonable request to the authors

Mikalena Xenophontos -, Rosie Ross -, Zoe Rutter-Locher –, Sarah Hill -, Sarah Ryan –, Mosab Ali Awadelkareem -, Shing T. Law -, David L. Bennett -, Christopher D. Buckley -, Frances Humby -, Bruce W. Kirkham -, Franziska Denk -, Leonie S. Taams -

## References

1. Shi G, Liao X, Lin Z, et al. Estimation of the global prevalence, incidence, years lived with disability of rheumatoid arthritis in 2019 and forecasted incidence in 2040: results from the Global Burden of Disease Study 2019. Clin Rheumatol 2023;42(9):2297–309. doi: 10.1007/s10067-023-06628-2 [published Online First: 20230609]

2. Vergne-Salle P, Pouplin S, Trouvin AP, et al. The burden of pain in rheumatoid arthritis: Impact of disease activity and psychological factors. Eur J Pain 2020;24(10):1979–89. doi: 10.1002/ejp.1651 [published Online First: 20200911]

3. Heiberg T, Kvien TK. Preferences for improved health examined in 1,024 patients with rheumatoid arthritis: pain has highest priority. Arthritis Rheum 2002;47(4):391–7. doi: 10.1002/art.10515

4. Gullick NJ, Ibrahim F, Scott IC, et al. Real world long-term impact of intensive treatment on disease activity, disability and health-related quality of life in rheumatoid arthritis. BMC Rheumatol 2019;3:6. doi: 10.1186/s41927-019-0054-y [published Online First: 20190225]

5. McWilliams DF, Ferguson E, Young A, et al. Discordant inflammation and pain in early and established rheumatoid arthritis: Latent Class Analysis of Early Rheumatoid Arthritis Network and British Society for Rheumatology Biologics Register data. Arthritis Res Ther 2016;18(1):295. doi: 10.1186/s13075-016-1186-8 [published Online First: 20161213]

6. Taylor P, Manger B, Alvaro-Gracia J, et al. Patient perceptions concerning pain management in the treatment of rheumatoid arthritis. J Int Med Res 2010;38(4):1213–24. doi: 10.1177/147323001003800402

7. Mapp PI, Kidd BL, Gibson SJ, et al. Substance P-, calcitonin gene-related peptide-and C-flanking peptide of neuropeptide Y-immunoreactive fibres are present in normal synovium but depleted in patients with rheumatoid arthritis. Neuroscience 1990;37(1):143–53. doi: 10.1016/0306-4522(90)90199-e

8. Pereira da Silva JA, Carmo-Fonseca M. Peptide containing nerves in human synovium: immunohistochemical evidence for decreased innervation in rheumatoid arthritis. J Rheumatol 1990;17(12):1592–9.

9. Vasconcelos DP, Jabangwe C, Lamghari M, et al. The Neuroimmune Interplay in Joint Pain: The Role of Macrophages. Front Immunol 2022;13:812962. doi: 10.3389/fimmu.2022.812962 [published Online First: 20220310]

10. Rutter-Locher Z, Kirkham BW, Bannister K, et al. An interdisciplinary perspective on peripheral drivers of pain in rheumatoid arthritis. Nature Reviews Rheumatology 2024 doi: 10.1038/s41584-024-01155-z

11. McWilliams DF, Dawson O, Young A, et al. Discrete Trajectories of Resolving and Persistent Pain in People With Rheumatoid Arthritis Despite Undergoing Treatment for Inflammation: Results From Three UK Cohorts. The Journal of Pain 2019;20(6):716–27. doi: 10.1016/j.jpain.2019.01.001

12. Woolf CJ. Central sensitization: implications for the diagnosis and treatment of pain. Pain 2011;152(3 Suppl):S2–S15. doi: 10.1016/j.pain.2010.09.030 [published Online First: 20101018]

13. Chaabo K, Chan E, Garrood T, et al. Pain sensitisation and joint inflammation in patients with active rheumatoid arthritis. RMD Open 2024;10(1) doi: 10.1136/rmdopen-2023-003784 [published Online First: 20240319]

14. Parsonage G, Filer AD, Haworth O, et al. A stromal address code defined by fibroblasts. Trends Immunol 2005;26(3):150–6. doi: 10.1016/j.it.2004.11.014

15. Buckley CD, Ospelt C, Gay S, et al. Author Correction: Location, location, location: how the tissue microenvironment affects inflammation in RA. Nat Rev Rheumatol 2021;17(4):246. doi: 10.1038/s41584-021-00602-5

16. Buckley CD, Filer A, Haworth O, et al. Defining a role for fibroblasts in the persistence of chronic inflammatory joint disease. Ann Rheum Dis 2004;63 Suppl 2(Suppl 2):ii92–ii95. doi: 10.1136/ard.2004.028332

17. Chakrabarti S, Hore Z, Pattison LA, et al. Sensitization of knee-innervating sensory neurons by tumor necrosis factor-alpha-activated fibroblast-like synoviocytes: an in vitro, coculture model of inflammatory pain. Pain 2020;161(9):2129–41. doi: 10.1097/j.pain.0000000000001890

18. Bai Z, Bartelo N, Aslam M, et al. Synovial fibroblast gene expression is associated with sensory nerve growth and pain in rheumatoid arthritis. Sci Transl Med 2024;16(742):eadk3506. doi: 10.1126/scitranslmed.adk3506 [published Online First: 20240410]

19. Wei K, Korsunsky I, Marshall JL, et al. Notch signalling drives synovial fibroblast identity and arthritis pathology. Nature 2020;582(7811):259–64. doi: 10.1038/s41586-020-2222-z [published Online First: 20200422]

20. Zhang F, Jonsson AH, Nathan A, et al. Deconstruction of rheumatoid arthritis synovium defines inflammatory subtypes. Nature 2023;623(7987):616–24. doi: 10.1038/s41586-023-06708-y

21. Zhang F, Wei K, Slowikowski K, et al. Defining inflammatory cell states in rheumatoid arthritis joint synovial tissues by integrating single-cell transcriptomics and mass cytometry. Nature Immunology 2019;20(7):928–42. doi: 10.1038/s41590-019-0378-1

22. Denk F, Bennett DL, McMahon SB. Nerve Growth Factor and Pain Mechanisms. Annu Rev Neurosci 2017;40:307–25. doi: 10.1146/annurev-neuro-072116-031121 [published Online First: 20170424]

23. Brenn D, Richter F, Schaible HG. Sensitization of unmyelinated sensory fibers of the joint nerve to mechanical stimuli by interleukin-6 in the rat: an inflammatory mechanism of joint pain. Arthritis Rheum 2007;56(1):351–9. doi: 10.1002/art.22282

24. Jang JH, Clark DJ, Li X, et al. Nociceptive sensitization by complement C5a and C3a in mouse. Pain 2010;148(2):343–52. doi: 10.1016/j.pain.2009.11.021 [published Online First: 20091223]

25. Spofford CM, Mohan S, Kang S, et al. Evaluation of leukemia inhibitory factor (LIF) in a rat model of postoperative pain. J Pain 2011;12(7):819–32. doi: 10.1016/j.jpain.2011.02.351

26. Li Y, Gray EH, Ross R, et al. Blockade of rheumatoid arthritis synovial fluid-induced sensory neuron activation by JAK inhibitors. bioRxiv 2024:2024.08.19.608613. doi: 10.1101/2024.08.19.608613

27. Croft AP, Campos J, Jansen K, et al. Distinct fibroblast subsets drive inflammation and damage in arthritis. Nature 2019;570(7760):246–51. doi: 10.1038/s41586-019-1263-7 [published Online First: 20190529]

28. Melzack R. The McGill Pain Questionnaire: major properties and scoring methods. Pain 1975;1(3):277–99. doi: 10.1016/0304-3959(75)90044-5

29. Rifbjerg-Madsen S, Christensen AW, Christensen R, et al. Pain and pain mechanisms in patients with inflammatory arthritis: A Danish nationwide cross-sectional DANBIO registry survey. PLoS One 2017;12(7):e0180014. doi: 10.1371/journal.pone.0180014 [published Online First: 20170707]

30. Cruz-Almeida Y, Mehta B, Haelterman NA, et al. Clinical and biobehavioral phenotypic assessments and data harmonization for the RE-JOIN research consortium: Recommendations for common data element selection. Neurobiol Pain 2024;16:100163. doi: 10.1016/j.ynpai.2024.100163 [published Online First: 20240822]

31. D’Agostino MA, Terslev L, Aegerter P, et al. Scoring ultrasound synovitis in rheumatoid arthritis: a EULAR-OMERACT ultrasound taskforce-Part 1: definition and development of a standardised, consensus-based scoring system. RMD Open 2017;3(1):e000428. doi: 10.1136/rmdopen-2016-000428 [published Online First: 20170711]

32. Donlin LT, Rao DA, Wei K, et al. Methods for high-dimensional analysis of cells dissociated from cryopreserved synovial tissue. Arthritis Res Ther 2018;20(1):139. doi: 10.1186/s13075-018-1631-y [published Online First: 20180711]

